# Transparency in epidemiological analyses of cohort data - A case study of the Norwegian Mother, Father, and Child cohort study (MoBa)

**DOI:** 10.1101/2024.12.05.24318481

**Authors:** Timo Roettger, Adrian Dahl Askelund, Viktoria Birkenæs, Ludvig Daae Bjørndal, Agata Bochynska, Bernt Damian Glaser, Tamara Kalandadze, Max Korbmacher, Ivana Malovic, Julien Mayor, Pravesh Parekh, Daniel S. Quintana, Laurie J. Hannigan

## Abstract

**Background:** Epidemiological research is central to our understanding of health and disease. Secondary analysis of cohort data is an important tool in epidemiological research but is vulnerable to practices that can reduce the validity and robustness of results. As such, adopting measures to increase the transparency and reproducibility of secondary data analysis is paramount to ensuring the robustness and usefulness of findings. The uptake of such practices has not yet been systematically assessed.

**Methods:** Using the Norwegian Mother, Father and Child Cohort study (MoBa; Magnus et al., 2006, 2016) as a case study, we assessed the prevalence of the following reproducible practices in publications between 2007-2023: preregistering secondary analyses, sharing of synthetic data, additional materials, and analysis scripts, conducting robustness checks, directly replicating previously published studies, declaring conflicts of interest and publishing publicly available versions of the paper.

**Results:** Preregistering secondary data analysis was only found in 0.4% of articles. No articles used synthetic data sets. Sharing practices of additional data (2.3%), additional materials (3.4%) and analysis scripts (4.2%) were rare. Several practices, including data and analysis sharing, preregistration and robustness checks became more frequent over time. Based on these assessments, we present a practical example for how researchers might improve transparency and reproducibility of their research.

**Conclusions:** The present assessment demonstrates that some reproducible practices are more common than others, with some practices being virtually absent. In line with a broader shift towards open science, we observed an increasing use of reproducible research practices in recent years. Nonetheless, the large amount of analytical flexibility offered by cohorts such as MoBa places additional responsibility on researchers to adopt such practices with urgency, to both ensure the robustness of their findings and earn the confidence of those using them. A particular focus in future efforts should be put on practices that help mitigating bias due to researcher degrees of freedom – namely, preregistration, transparent sharing of analysis scripts, and robustness checks. We demonstrate by example that challenges in implementing reproducible research practices in analysis of secondary cohort data - even including those associated with data sharing - can be meaningfully overcome.

## 1. Background

Epidemiological research is central to our understanding of why and how diseases and disorders originate and spread over time. For example, the Norwegian Mother, Father and Child Cohort Study (MoBa; Magnus et al., 2006) was created to investigate diseases and disorders, collecting longitudinal information about almost 100,000 mothers alongside their children and their children’s fathers over the last 25 years. This dataset has allowed researchers to successfully answer questions about fertility, cognitive development and the development of the human brain, among many other research areas (Magnus et al., 2016).

### 1.1 Epidemiological research and the garden of forking paths

Epidemiological data sets, such as MoBa, are rich and complex, containing hundreds of variables, and offer a myriad of ways to learn from them. To analyze these datasets appropriately, many different methodological and analytical choices need to be made, all of which may influence the final interpretation of the data (Wicherts et al., 2016). This range of analytical decisions introduces so-called researcher degrees of freedom. On the one hand, they represent opportunities to look at the data set from many different angles, which, in turn, allows researchers to make important discoveries and generate novel hypotheses (e.g., Box, 1976; De Groot, 2014; Tukey, 1977). On the other hand, idiosyncratic choices can lead to categorically different interpretations (Simmons et al., 2011). Recent studies have shown that independent data analysts analyze the same data set in vastly different ways, leading them to arrive at different conclusions when trying to answer the same research question (e.g., Silberzahn et al., 2018; Dutilh et al., 2019; Starns et al., 2019; Coretta et al., 2023).

For example, Silberzahn et al. (2018) invited 29 independent research teams to test the same hypothesis: whether soccer referees are more likely to issue red cards to players with darker skin tones compared to those with lighter skin tones. The teams employed a wide range of analytical approaches, leading to varying results - 20 teams (69%) found evidence supporting the hypothesis, while 9 teams (31%) did not. Among the 29 analyses, there were 21 distinct combinations of predictors used. Notably, the diversity in outcomes could not be explained by the researchers’ prior beliefs about the topic or by peer evaluations of the quality of their analyses. The findings highlight that variability in analytic results may be an inherent feature of the scientific process, rather than a reflection of bias or differences in expertise. Follow up studies further showed that even seemingly inconsequential choices regarding data cleaning, variable operationalisation, and model structure can lead to categorically different conclusions (e.g. Coretta et al., 2023).

With rich cohort data such as MoBa, the “garden of forking paths” of possible analytical pipelines (Gelman & Loken, 2014) is so vast - and the amount of researcher degrees of freedom is so large - that arguably any hypothesis can be supported by the data if the data are interrogated often enough. Indeed, some recent empirical efforts to either replicate, reproduce, or generalise epidemiological findings have been unsuccessful (Border et al., 2019; Seibold et al., 2021; Orben et al., 2019). This underlines the urgency of addressing these issues in research using secondary data analysis.

### 1.2 Transparent scientific practices

Under the broad umbrella of “Open Science”, a number of reform efforts have recently been proposed across disciplines (e.g. Kidwell et al., 2016; Klein et al., 2018; Zwaan et al., 2018; Korbmacher et al., 2023). The main focus of these efforts has been to increase the transparency of the research process and its outputs by, for example, establishing and archiving a public version of a preregistered research plan (Nosek et al., 2018), and sharing of research materials and procedures, raw and processed data, and analysis scripts (Gilmore et al., 2018; Lindsay, 2017). The central goal of these efforts is to allow the scientific community and often also the broader public to access relevant information so that they can critically evaluate scientific claims (Munafò et al., 2017; Vazire, 2017). Some of these practices directly tackle challenges related to analytical flexibility. Sharing statistical protocols and data enables transparent and unambiguous communication of all relevant analytical decisions and, in turn, allows other researchers to assess the robustness of an analysis against other possible analyses. Preregistration draws a transparent line between initially planned analyses and post hoc exploratory analyses (Nosek et al., 2018), makes questionable research practices such as selective reporting and hypothesizing after results are known detectable (John et al., 2012), and helps reduce researcher degrees of freedom (Mertzen et al., 2021; Simmons et al., 2011; Wicherts et al., 2016), making it a particularly useful tool in epidemiological research.

Beyond directly addressing concerns related to researcher degrees of freedom, transparent practices promise general benefits for both individual researchers and science at large. More transparent research can accelerate discovery by allowing re-use of research materials, procedures and data, which can in turn facilitate collaborations (Boland et al., 2017), and increase efficiency and sustainability (Lowndes et al., 2017). Transparent sharing of research materials, data and scripts makes it easier to detect and correct errors (Nuijten et al., 2016). It can also aid in facilitating evidence synthesis (e.g. meta-analyses; Pigott & Polanin, 2020) and enable independent reproductions and replications of results (Munafò et al., 2017). Given this propagation of the effects of transparent and reproducible practices across all levels of the research process – and that the goal of all epidemiological research is to benefit patients and their families – their implementation can ultimately be seen as a means of achieving this goal by enhancing and accelerating the clinical impact of findings.

### 1.3 The present paper

Recent assessments of biomedicine (Wallach et al., 2018), social sciences (Hardwicke et al., 2020), psychological sciences (Hardwicke et al., 2022) and language sciences (Bochynska et al., 2023) suggest that rates of transparent practices are still quite low. In epidemiological analyses of cohort data, researchers may perceive specific challenges to implementing transparent practices (Baldwin et al., 2022; van den Akker et al., 2019; Weston et al., 2019). For example, epidemiological cohorts often contain sensitive and/or personal data that cannot be shared openly. Even sharing derivatives of the original data can contain identifying information (Rocher et al., 2019). Thus, the aim of the current paper is to assess the current level of transparent practices found in secondary analysis of cohort data and to offer concrete and actionable advice on how to be as transparent as possible given the existing limitations and challenges. First, we aim to assess and quantify the adoption of transparent practices in secondary data analysis of cohort data using MoBa as a case study. Second, we will identify and demonstrate solutions that can facilitate the adoption of transparency practices in cohort studies, with a particular focus on practices that tackle issues related to researcher degrees of freedom.

## 2. Methods

### 2.1 Assessing transparent research practices in MoBa

As a case study, we assessed reproducible and transparent practices in secondary data analysis using the MoBa dataset. We chose MoBa for several reasons: First, the MoBa cohort has resulted in more than thousand research articles, thus providing a representative snapshot of scientific practices while the sample size remains feasible for a systematic meta scientific assessment. Second, MoBa-based research has been published across a long enough time span (>17 years) to meaningfully investigate trends over time. Third, data collection for MoBa is ongoing and thus likely to result in many more future publications. Assessing research on MoBa, thus, gives us the opportunity to encourage researchers engaging with MoBa to adopt more open and reproducible practices.

Specifically, we looked at the implementation of the following practices which are currently considered among best practices for open and credible research: preregistering secondary analyses, sharing of synthetic data, sharing of meta-data and summary statistics, sharing of additional materials, including analysis scripts, conducting robustness checks, directly replicating previously published studies, declaring conflicts of interest and publishing of publicly available versions of the paper. Our study’s design and data collection plan was uploaded as a preregistration prior to data collection (https://osf.io/ef4t5/). All materials, data, and analysis scripts related to this study are publicly available on Open Science Framework: https://osf.io/2jqxv/.

#### 2.1.1 Sample

As the primary data source, we retrieved the list of publications that have used the MoBa dataset from the MoBa website^1^ and obtained 1005 articles. After a pilot evaluation of 60 randomly sampled entries, the remaining 945 articles were coded by six raters.

#### 2.1.2 Procedure

Measured variables and their operationalisations are shown in Table 1. We coded the year of publication and the type of study by examining title, abstract, and if necessary, the methods section, to establish the study characteristics. We coded whether authors claimed the presence of preregistrations, synthetic data, other non-primary data such as metadata and summary statistics, analysis protocols or additional materials, by examining relevant parts of the method and result section of each article. If one of these aspects was present, we noted how this information was accessible, and whether we could indeed access it or not. We moreover assessed whether the authors claimed to have replicated a previous study or not and whether authors declared the absence or presence of a conflict of interest. Data extraction was executed via a Google Form consisting of questions to the coder and response options, see https://osf.io/47xmh.

**Table 1:**
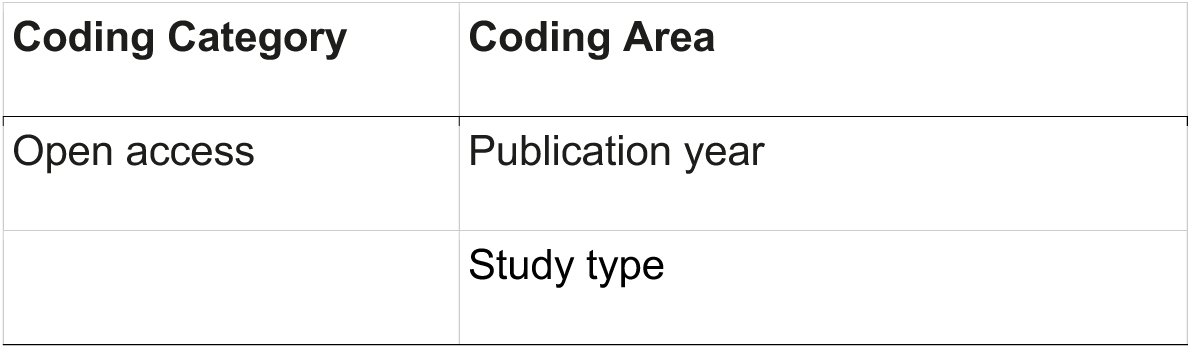
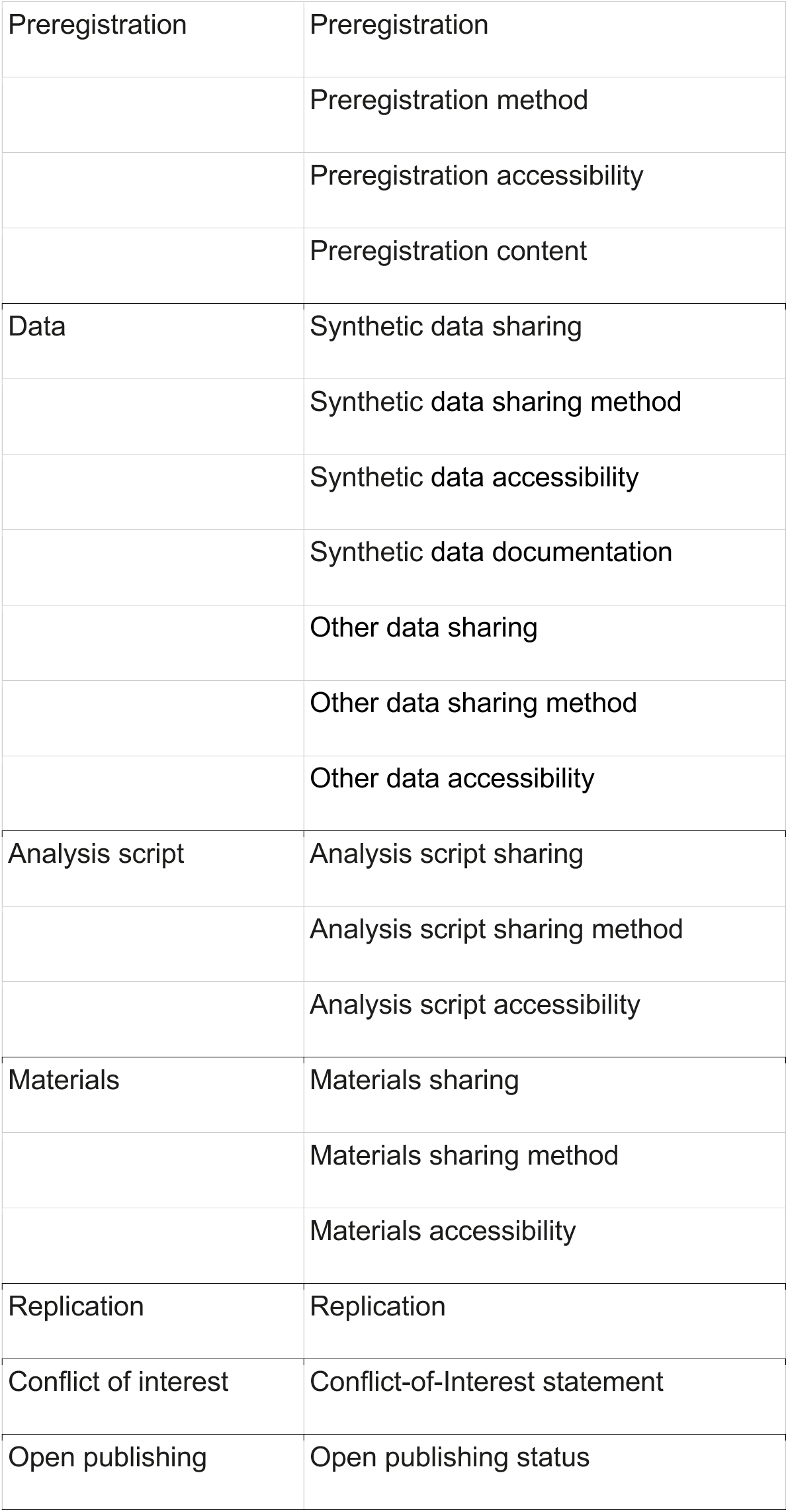
Main measured variables.

Each coder assessed individual articles for each measured variable, recording outcomes in a structured form (https://osf.io/2qdzb). In total, 80% of the 945 target articles were randomly drawn and coded by single coders. The remaining subset (20%) was coded by two coders to assess inter-rater reliability. See the preregistration for more details on the coding procedure.

### 2.2 How to be as transparent as possible - a toy example

We also aim to demonstrate a worked example of how researchers analyzing MoBa data can use current tools and standards for transparent practices in conducting and reporting their work. To that end, we simulated data based on the characteristics of real MoBa data in order to present a realistic example of a reproducibly planned and reported MoBa analysis. The key features that we emphasise in this example are preregistration, detailed reporting of deviations and unregistered steps, robustness checks, sharing of well-annotated analytic code, and sharing of synthetic data to facilitate computational reproducibility. All materials are available (https://osf.io/nm9ej/) and can be used as points of departure for future efforts.

In our hypothetical research project, researchers were interested in testing the independent effects of age (*M* = 3.46 years, *SD* = 2.76) and breastfeeding duration (*M* = 5.47 months, *SD* = 3.73) on height measured on six occasions during childhood in MoBa and assessing whether these effects vary according to sex (a categorical variable with levels: “Female” and “Male”).

## 3. Results

### 3.1 Assessment of MoBa

Coders agreed for 91% of all items. Discrepancies between the first and the second coder were not obviously linked to any particular coding category and were easily resolved through discussion. The following overview is based on the consensus ratings. Of the 943 articles, 799 (84.7%) were coded as in-principle accessible. Only those articles were further analysed. 58 articles could not be accessed by the coder (6.2%); 24 were coded as not being a full article, e.g. only a conference abstract or an erratum (2.5%); two were coded as not being written in English (0.2%); and 60 articles were coded as having other issues, including papers that turned out to actually not analyze MoBa (6.4%).

Of those articles that were in principle accessible, 733 articles were coded as presenting empirical data and using the MoBa corpus (91.7%). All rates reported below refer to these 733 articles. Figure 1 summarises the results in relation to the articles’ year of publication. Green represents transparent practices, and gray represents non-transparent practices. Individual assessment categories are discussed below. A more detailed presentation of data is accessible here (https://osf.io/ec6tf).

**Figure 1:**
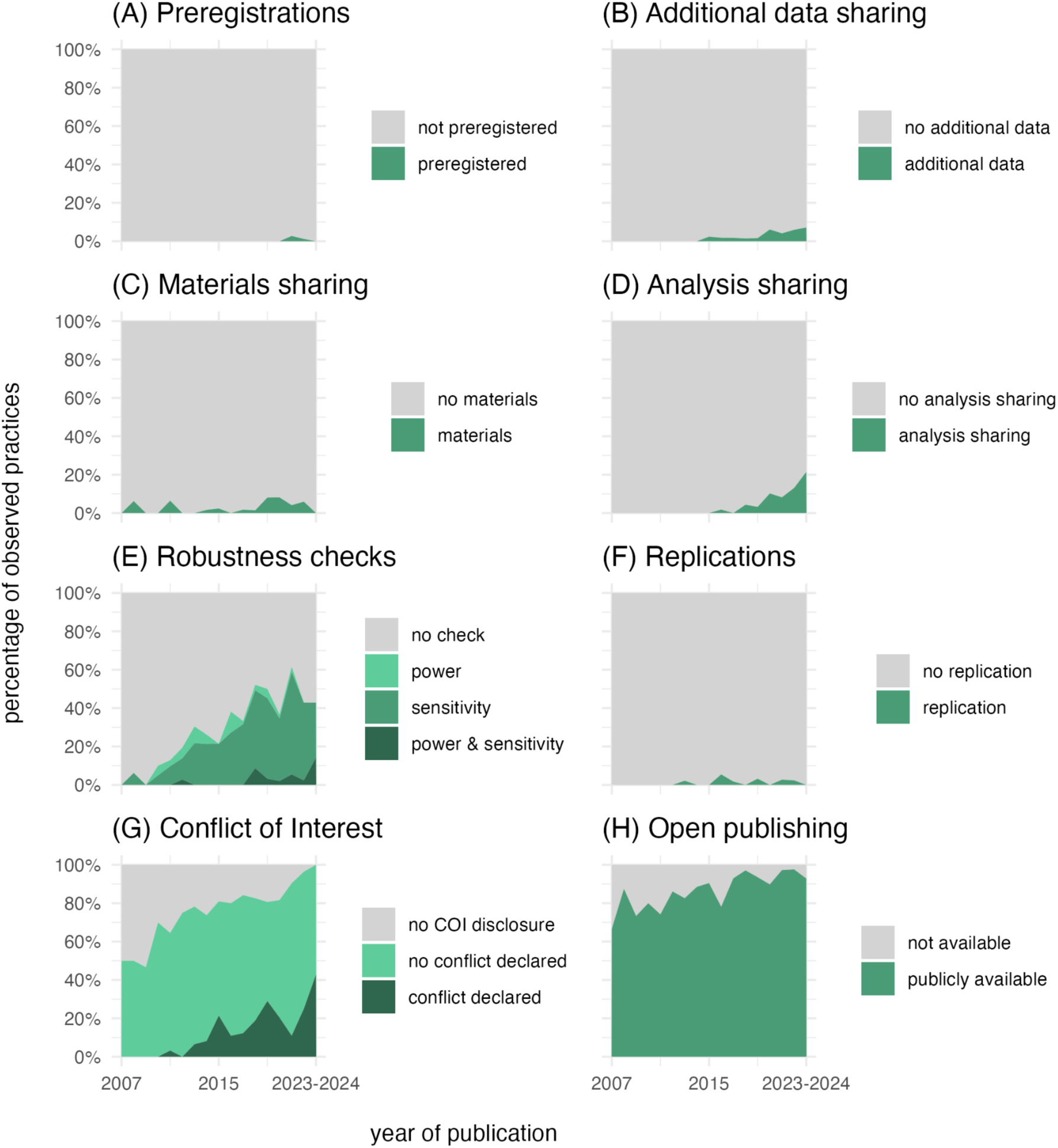
Percentage of transparent practices for articles as a function of publication year (2007-2024). Year 2023 and 2024 were collapsed due to a small number of *n*.

#### 3.1.1 Preregistration, data and analysis sharing

Despite its relevance for secondary data analysis, there were only 3 articles in the entire sample (0.4%) that reported having preregistered aspects of their analysis. None of the articles reported having used synthetic data.

In our corpus, we found 17 articles to share resources that could be considered some form of non-primary data (2.3%), which mostly referred to additional metadata, validation data, or summary statistics. However, only 31 of assessed articles were coded as reporting to share some form of analysis protocols either in form of step-by-step instructions or code (4.2%).

Despite these low rates, there is a trend towards more transparent practices: both additional data sharing, and analysis sharing indicate a trend upwards with 7.1% of articles sharing additional data and 21.4% of articles sharing their analysis in 2023/2024. Both practices were non-existent in the first half of the publication range (see Fig. 1B and 1D).

#### 3.1.2 Robustness checks

Given the analytical flexibility in secondary data analysis, it is useful to evaluate how robust a statistical outcome is under different analytical assumptions. Such sensitivity analyses were indeed not uncommon in our sample: one third of articles stated that some form of a sensitivity analysis was performed (239 articles, 33%). 44 of assessed articles reported a power analysis (6%). Given the lack of sharing practices of scripts and non-primary data, the exact nature of both these sensitivity and power analyses often remained unclear. While the overall rates for robustness checks were low, general upward trends were apparent with 42.9% of articles performing a sensitivity analysis and 14.3% a power analysis in 2023/2024, both of which were almost non-existent at the beginning of the publication range (see Fig. 1E).

#### 3.1.3 Other transparency practices

In our assessment, 24 articles (3.4%) reported to sharing some form of additional materials such as algorithmic description of quality control procedures (e.g., Isungset et al., 2022) or insights into the selection of items and results from measurement models (Øksendal et al., 2022). 11 articles reported to have directly replicated a previous study (1.5%). Compared to other fields, MoBa articles have a high rate of declaring conflicts of interest, approaching ceiling effects for recently published articles. Only 142 of assessed articles were coded as not reporting a conflict-of-interest statement (19.3%); 488 reported no conflict of interest (66.2%); and 107 reported an existing conflict of interest (14.5%). It is important to note that these rates likely do not reflect voluntary practices of researchers but reflect how many journals require this form of disclosure. When it comes to publicly sharing a version of the manuscript, 661 of assessed articles were publicly available according to our definition (89.7%) which is a much higher rate than similar assessments in other fields (e.g., biomedicine: 25%; Wallach et al., 2018).

### 3.2 Demonstrating transparent practices

#### 3.2.1 Preregistering the analysis and document deviations

Here we present a full toy example that demonstrates workflows and tools that can be used to increase transparency for secondary data analyses of cohort data. The preregistration of the toy example is uploaded to the Open Science Framework Registry prior to accessing the data for analyses and can be accessed here: https://osf.io/eczpq. The hypothetical researchers investigate relationships between age, breastfeeding duration, and childhood height in MoBa and aim to limit researcher degrees of freedom in the specific context of analysing pre-existing data through a thorough preregistration. In particular, this includes specification of inclusion/exclusion criteria, operationalisation of relevant variables that will be included in analytic models, and specification of hypotheses that will be tested alongside the applied inference criteria. The researchers also disclose the nature and extent of their knowledge of MoBa data from prior work.

The specific variables of our toy example were chosen as they illustrate variables that are typically used in MoBa. For instance, questionnaire data may look and act differently than what was expected in the preregistration phase. We emulated this here by simulating irregular distributions for mother-reported breastfeeding duration, with large idiosyncratic asymmetries in its distribution (as well as missing data), suggesting at least three underlying data generating patterns for the variable. Given these irregularities, in the analysis phase, the researchers decide to not treat the variable as a continuous predictor as preregistered but rather as a categorical predictor that bins breastfeeding into short (1-2 months), medium (3-5 months), and long duration (>5 months). Moreover, when cleaning the height variables, the hypothetical researchers realise that the preregistration was insufficiently precise about how “valid responses” should be defined and therefore implement a +/- 3 SD threshold for outlier removal. These issues will likely be familiar to readers accustomed to performing secondary data analyses and may even be part of the explanation for the limited uptake of preregistration for this kind of work in epidemiological studies. However, there are easy-to-implement solutions for such situations.

In our example, the researchers’ note their deviations and unregistered steps in a specially formatted table (Tab. 2, adapted from Willroth et al. 2024), designed to ensure transparency for readers. In implementing their deviation (categorising the breastfeeding variable), they are mindful of maintaining their Type 1 error rate at the prespecified level (introducing a single likelihood ratio test to assess the effect of breastfeeding, which would otherwise - with the exposure now a categorical variable - correspond to multiple model parameters). To interrogate the impact of their modified approach on their results, they additionally perform a sensitivity analysis with a slightly different categorisation of the breastfeeding duration variable (0-5 months as “early cessation”, 6 months as “recommended”, and over 6 months as “late cessation”). Likewise, when implementing their unregistered step (operationalising “valid responses” on the measure of height as those within 3 standard deviations of the mean), they again perform an additional sensitivity analysis (using a +/- 2 SD threshold) to interrogate the impact of this decision on the results and communicate this to readers.

**Table 2:** Preregistration Deviations Table for the Toy Example (the template can be accessed <u>here</u>)

| Details |  | Original Wording | Deviation Description | Reader Impact |
| --- | --- | --- | --- | --- |
| Type | Data Preparation | Section 3.1 of preregistration: “Breastfeeding duration: a continuous variable measured in months, derived based on mothers’ reports of breastfeeding in the six and 18 month questionnaires.” | Based on the distribution of the breastfeeding variable, this variable was categorised into “None/early cessation”, “Up to 6 months”, and “Extended duration”. The variable is not treated as ordinal due to the possibility of qualitative differences between non-/early stopping breastfeeders and others in the sample. | The hypothesis tests regarding breastfeeding duration (H2 and H4) are now evaluated based on a likelihood ratio test to compare the fit of the model with and without this variable. Adopting this approach, rather than drawing inferences based on beta coefficients for contrasts, ensures that the alpha for these hypothesis tests remains at 0.05. This deviation should have a limited impact on readers’ interpretation of results. The impact, on the results, of the specific implementation of this deviation are examined in sensitivity analyses (wherein an alternative categorisation of the breastfeeding variable is included). |
| Reason | New knowledge |  |  |  |
| Timing | After data access |  |  |  |

Unregistered Steps
| Details |  | Original Wording | Unregistered Step Description | Reader Impact |
| --- | --- | --- | --- | --- |
| Type | Data Preparation | In the preregistration, we stated that we would “retain all valid responses” in section 3.5 Statistical outliers. | In order to operationalise “valid responses”, we set height values $\geq 3$ standard deviations from the mean to missing. | This unregistered step was an omission of detail in the preregistration and should have a limited impact on readers’ interpretation of the results. The impact, on the results, of the specific |
| Timing | After results known |  |  |  |
| | | | | implementation of this step are examined in sensitivity analyses (wherein a threshold of $\geq 2$ standard deviations is used). |

#### 3.2.2 Sharing of analytic code and metadata

After completing the analyses, the researchers report all the details of their data preparation and analyses in the methods section of their manuscript – with much of the detail imported from their preregistration. However, they are aware that such a report, no matter how diligently compiled, is unlikely to be sufficiently detailed to facilitate reproducibility without the statistical protocol or code used to implement these steps. They provide analysis scripts in an online repository (https://osf.io/nm9ej/), organised so that its contents are easy to navigate and understand to possible users. A readme file in the repository itemises and describes the files it contains, and analytic scripts include comments to allow readers to follow them and relate their contents back to the manuscript, and the results obtained by the researchers.

#### 3.2.3 Create synthetic data to allow reproducibility

The researchers wish to share their data to allow others to directly reproduce their findings. However, the consent given by MoBa participants does not allow data to be shared openly. Therefore, they create a synthetic dataset which preserves the quantitative relationships between variables without including any information corresponding to real individuals from the sample. This dataset, available in a designated data repository (https://osf.io/nm9ej/), can be downloaded for use with the analytic code shared, allowing the code to be run and the results reproduced. It is important to note however that synthetic data do not allow for an exact reproduction of the original data. General quantitative relationships are maintained but exact values are slightly different since the involved data simulation process is stochastic. While sharing of synthetic data based on secondary analysis of cohorts like MoBa is relatively new and subject to some specific considerations (Jordon et al., 2022; Major-Smith et al., 2024), guidance exists to help researchers make use of this important tool for reproducibility (e.g., Quintana, 2020). For our example, the online repository includes code used to (a) generate synthetic data using the *synthpop* package in R (Nowok et al., 2016), (b) verify that it does not disclose information about individuals in the real dataset, and (c) validates that it produces similar results to the original (simulated) data. Admittedly, currently available materials such as our own use rather simple example data. Depending on the analysis and the data, creating accurate and appropriate synthetic data might be challenging for complex data sets and might require particular statistical expertise. However, we believe it is important to emphasise that even an imperfect attempt to make research more reproducible is worthwhile, commendable and more useful than not attempting it at all.

## 4. Discussion

Our assessment of transparent practices in MoBa showed that some transparent practices in MoBa are generally low. In line with assessments from other fields (Hardwicke et al., 2020; 2022; Rowhani-Farid & Barnett, 2018; Wallach et al., 2018; Bochynska et al., 2023), preregistration, sharing of additional data, additional materials and analysis script were rare, although some of them indicated a positive trend towards higher rates in recent years.

Preregistration is still not a common practice in secondary data analysis of the MoBa cohort. Preregistration can be perceived as challenging due to researchers’ prior knowledge of the data, or the perception that preregistrations are inappropriate because analytical decisions need to be made contingent on properties of the data. As illustrated by our demonstration, all of these perceived challenges have straightforward and actionable solutions (Baldwin et al., 2022): Researchers can preregister their plan (alongside declarations of their prior knowledge about the data) either by using templates specific to secondary data analysis provided on websites, such as the Open Science Framework Registry (https://osf.io/registries) and AsPredicted (https://aspredicted.org/), or by publicly archiving their preregistration in other formats. Necessary deviations can be accommodated and recorded in ways that preserve false discovery rates at pre-registered levels and prioritise analytical transparency. Additionally, more and more journals offer specific article types called Registered Reports, which are peer-reviewed preregistrations (Nosek & Lakens, 2014)^2^. Even though the uptake of this format remains still rather limited for medical and epidemiological journals, there are exceptions (e.g., *BMC Medicine*, *Nature Human Behavior*).

Sharing practices of additional data, additional materials, and analysis scripts were also low in our assessment. Raw data of MoBa can categorically not be shared because it is outside of participants’ consent and any form of derived data tables might also not easily be shared due to legal restrictions and privacy concerns. A possible way to circumvent these issues is the generation of synthetic data (Quintana, 2020, Major-Smith et al. 2024), i.e. simulated data that retain statistical properties of the original data but remove all identifying information. None of the articles reported using synthetic data, which in principle would have allowed the reader to reproduce their analysis without violating data protection laws. However, it is important to note that this is a relatively new approach for sharing sensitive data and thus not widely known. To raise awareness and reassure both administrators and participants of cohort studies about the appropriateness of this approach, we offered a concrete example of synthetic data simulation in our demonstration. Sharing synthetic datasets for analyses such as those performed in our example can be transformative for the field. The availability of tailored R packages (such as *synthpop*) means that, in many cases, the creation of synthetic data can be both a computationally and practically simple step to implement at the end of an analytic pipeline. For more complex designs that exceed the restrictions of available software, manual simulations assuming simple distributions can often offer an acceptable approximation. Even in these "higher cost scenarios", we argue that the benefits outweigh the investments, as shared synthetic data has the potential to improve the quality and efficiency of the peer review process, empower readers to understand and make secondary use of analytic code, and facilitate the first phase of attempts to reproduce or interrogate analytic findings.

Complementary to synthetic data, sharing additional data such as metadata (i.e., information about the data) increases the findability, the transparency, and the usability of aspects of an analysis. Certain descriptive statistics allow researchers to reverse-engineer at least some statistical properties of numerical distributions. Despite these benefits, sharing these forms of information has not been widely adopted for MoBa research. The repository associated with our demonstration can be used as a model for ways to enrich shared resources with appropriate metadata.

Whether additional or synthetic data is available or not, a step-by-step description of the analysis in the form of a script is the minimal requirement for other researchers to reproduce the results and evaluate how researchers arrived at their conclusions. While this practice was by no means a widespread standard throughout the earlier years of the period from which we assessed studies, it has become an expectation - and often mandate - of many publishers and funders in recent years. Yet, our assessment indicates that sharing practices of data analysis protocols are still not common. Sharing such resources, however, is important because humans are error-prone (Nuijten et al., 2016), and methodological descriptions often lack the detail to recreate or critically evaluate the data analysis pipeline (Hardwicke et al., 2018). The latter is a problem, because the outcome and interpretation of analyses has been shown to be dependent on analytical details (e.g., Silberzahn et al., 2018; Dutilh et al., 2019; Starns et al., 2019; Coretta et al., 2023).

The robustness of the outcome and interpretation of an analysis can be assessed by probing the sensitivity of the results to researcher degrees of freedom. Our demonstration offered simple sensitivity checks which can straightforwardly be scaled up. Particularly comprehensive versions of sensitivity analyses are so-called multiverse analyses (Steegen et al., 2016), which involves identifying and implementing all potential analytic choices that could justifiably be made to address the research question. These analyses often involve thousands of analytical pipelines, which may be impractical for some computationally intensive analyses of large-scale datasets. Nonetheless, sensitivity analyses at some scale are an important element of the toolbox for conducting secondary data analysis transparently and robustly.

Not surprisingly, power analyses are not very common for MoBa studies. Given that most inferential analyses in research on cohort data are based on versions of the null hypothesis significance testing framework, decision procedures are usually binary (i.e. is a relationship significant or not). A non-significant result can either reflect a true negative, i.e. the true absence of a relationship, or a false negative because the analysis lacked sufficient statistical power. To interpret a non-significant result in light of its false negative rate, analysts can a priori estimate the statistical power of a statistical test. However, power analysis is challenging for secondary data analysis and arguably of limited use because researchers have no control over the sample size or the measurement process. Researchers performing secondary data analysis can give readers of their research the best information to interpret their results in light of statistical power by ensuring that test results are supplemented with the presentation of estimates of effect sizes and corresponding indices of precision.

## 5. Conclusion

We demonstrated that some transparent practices in the Norwegian Mother, Father and Child Cohort study (MoBa), are more common than others, with some practices being virtually absent. It is important to acknowledge that our review assessed research published across a span of 17 years against criteria based on *present day* best practices, not all of which have been widely popularised for long. Nonetheless, even recent upward trends would – if they continue – be insufficient to see a majority of researchers analyzing MoBa data as reproducibly as possible within the next decade. The complexity of cohort corpora such as MoBa invites a large amount of analytical flexibility. If left unchecked, these researcher degrees of freedom, which can be either implicit or explicit, can facilitate bias and a proliferation of overconfident claims (Simmons et al., 2011; Gelman & Loken, 2014). A particular focus in future efforts in the field should be put on practices that help mitigating bias due to researcher degrees of freedom – namely, preregistration, transparent sharing of analysis scripts, and robustness checks.

With our toy example of a transparently reported MoBa analysis, we demonstrate that researchers performing secondary analyses of cohort data have no cause to be reluctant to adopt transparent and reproducible practices going forward. While our demonstration and its online materials illustrate just one possible way to document and transparently share analysis protocols, researchers without inclination to develop their own alternatives may consider using them as templates in future work.

Despite the general low rate of transparent and reproducible practices in our assessment, it is important to note that our assessment shows several positive trends over time. To us, these trends indicate that researchers are becoming more aware of the importance, usefulness, and feasibility of transparent practices. All in all, the field of secondary data analyses on cohorts in particular, and epidemiology in general seems to be moving towards a more transparent and reproducible future.

The adoption of transparent principles in the field of epidemiology can be further facilitated by making it possible and easy to register, share, and publish research outputs. Stakeholders such as funders, journals, and institutions should invest in and create infrastructures for these practices, and either incentivise them as a currency that increases the competitiveness of researchers or make them obligations when publishing, acquire funding, and compete for professional positions. We hope the present paper helps in tracking progress over time in secondary analysis of cohort data and epidemiology more broadly, enabling cross-disciplinary comparisons, and, through our accompanying materials, represent a useful point of departure for future efforts.

## Acknowledgment

Not applicable.

## Clinical trial number

Not applicable.

## Funding

TR was supported by the Research Council of Norway (ICONIC; #350026); PP was supported by the European Union’s Horizon 2020 research and innovation programme under the Marie Skłodowska-Curie grant (#801133) and by Research Council of Norway (#324252); ADA was supported by the South-Eastern Norway Regional Health Authority (#2020023) and the European Union’s Horizon Europe Research and Innovation programme (FAMILY; #101057529); BG was supported by the Research Council of Norway (#336085); DQ was supported by the Research Council of Norway (#324783) and the Kavli Trust; LH was supported by the South Eastern Regional Health Authority of Norway (Helse Sør-Øst; #2022083, #2019097).

## Author contributions

Authors contributed with the following roles to the present research article (following Contributor Role Taxonomy (CRediT):

**TR**: Conceptualtion, Data Curation, Software, Formal Analysis, Project Administration (lead), Supervision, Funding acquisition, Validation, Visualization, Methodology, Writing - original draft (lead), Writing - review and editing

**ADA**.: Conceptualization, Data Curation, Software, Formal Analysis, Funding acquisition, Methodology, Writing - original draft (support), Writing - review and editing

**VB**: Conceptualization, Methodology, Writing - original draft (support), Writing - review and editing

**LDB**: Conceptualization, Formal Analysis, Methodology, Writing - original draft (support), Writing - review and editing

**AB**: Conceptualization, Data Curation, Methodology, Writing - original draft (support), Writing - - review and editing

**BDG**: Conceptualization, Software, Formal Analysis, Validation, Methodology, Writing - original draft (support), Writing - review and editing

**TK**: Conceptualization, Data Curation, Methodology, Writing - original draft (support), Writing - review and editing

**MK**: Conceptualization, Data Curation, Software, Methodology, Writing - original draft (support), Writing - review and editing

**IM**: Conceptualization, Data Curation, Methodology, Writing - original draft (support), Writing - review and editing

**JM**: Conceptualization, Data Curation, Methodology, Writing - original draft (support), Writing - review and editing

**PP**: Conceptualization, Data Curation, Software, Formal Analysis, Funding acquisition, Validation, Methodology, Writing - original draft (support), Writing - review and editing

**DSQ**: Conceptualization, Formal Analysis, Methodology, Writing - original draft (support), Writing - review and editing

**LJH**: Conceptualization, Software, Formal Analysis, Project Administration (support), Funding acquisition, Validation, Methodology, Writing - original draft (support), Writing - review and editing

## Ethic approval and consent to participate

Not applicable.

## Consent for publication

We confirm that all authors of the manuscript have read and agreed to its content and are accountable for all aspects of the accuracy and integrity of the manuscript. No further consent requirement is applicable.

## Competing interests

AB, DQ, IM, JM, PP, TK and TR have engaged with meta-scientific research and have been actively involved in methodological debates about scientific practices, including transparency across disciplines. ADA, LDB, LH, and PP have published secondary analyses of the MoBa cohort. All other authors have no competing interests.

## Data availability statement

All dataset(s) and materials supporting the conclusions of this article are available in the OSF repository: https://osf.io/2jqxv/.

## Footnotes

1 https://www.fhi.no/en/ch/studies/moba/for-forskere-artikler/publications/

2 Since conducting our MoBa assessment, at least one registered report on MoBa data was published (Askelund et al. 2024), albeit by authors of the present paper only. The authors used a new wave of data collection in MoBa for the analyses to ensure that no prior knowledge of the data could influence the study design, a way to strictly control bias in registered reports based on cohort studies.

## Notes

### Summary of Updates

revised to add clarifications, amend typos, and streamline arguments

